# Determinants of SARS-CoV-2 infection in Italian healthcare workers: a multicenter study

**DOI:** 10.1101/2020.07.29.20158717

**Authors:** Paolo Boffetta, Francesco Violante, Paolo Durando, Giuseppe De Palma, Enrico Pira, Luigi Vimercati, Alfonso Cristaudo, Giancarlo Icardi, Emma Sala, Maurizio Coggiola, Silvio Tafuri, Vittorio Gattini, Pietro Apostoli, Giovanna Spatari, Working Group on SARS-CoV-2 infection in Italian healthcare workers.

## Abstract

**Background:** Healthcare workers (HCW) are at increased risk of being infected with SARS-CoV-2, yet limited information is available on risk factors of infection.

**Methods:** We pooled data on occupational surveillance of 10,654 HCW who were tested for SARS-CoV-2 infection in six Italian centers. Information was available on demographics, job title, department of employment, source of exposure, use of personal protective equipment (PPE), and COVID-19-related symptoms. We fitted multivariable logistic regression models to calculate odds ratios (OR) and 95% confidence intervals (CI).

**Findings:** The prevalence of infection varied across centers and ranged from 3.0% to 22.0%, being strongly correlated with that of the respective areas. Women were at lower risk of infection compared to men. Fever, cough, dyspnea and malaise were the symptoms most strongly associated with infection, together with anosmia and ageusia. No differences in the risk of infection were detected between job titles, or working in a COVID-19 designated department. Reported contact with a patient inside or outside the workplace was a risk factor. Use of a mask was strongly protective against risk of infection as was use of gloves. The use of a mask by the source of exposure (patient or colleague) had an independent effect in reducing infection risk.

## Introduction

Healthcare workers (HCW) are a group at high risk of infection in general^1^ and specifically SARS-CoV-2 infection.^2,3^ However, few studies have been reported in the literature on prevalence of COVID-19, and on related risk factors in this group of workers:^4^a study of 1654 HCW from England,^5^ one on 72 infected HCW from China,^6^ and two small studies of HCW from Switzerland and Singapore who had a contact with a case.^7,8^ Given the lack of information on determinants of infection in this important occupational group, and the relevance of such data for other groups of the population, we undertook an analysis of clinical and occupational data collected among more than 10,000 Italian HCW who were tested for presence of SARS-CoV-2 during March and April 2020. The project was conducted in seven academic centers under the auspices of the Scientific Committee of the Italian Society of Occupational Medicine.

## Results

A total of 10,654 HCW were included in the analysis. Period of testing, incidence and mortality rates of COVID-19 infection in the general population of the study areas are shown in Table 1. Key characteristics of the study population are included in Table 2. Women represented two thirds of HCW included in the analysis (which reflects the HCW demographics in Italy); average age ranged from 34 to 47 years, with overall mean of 45.4 years (sd 0.53). In all centers, nurses and doctors represented two thirds or more of HCW tested for SARS-CoV-2.

**Table 1.**
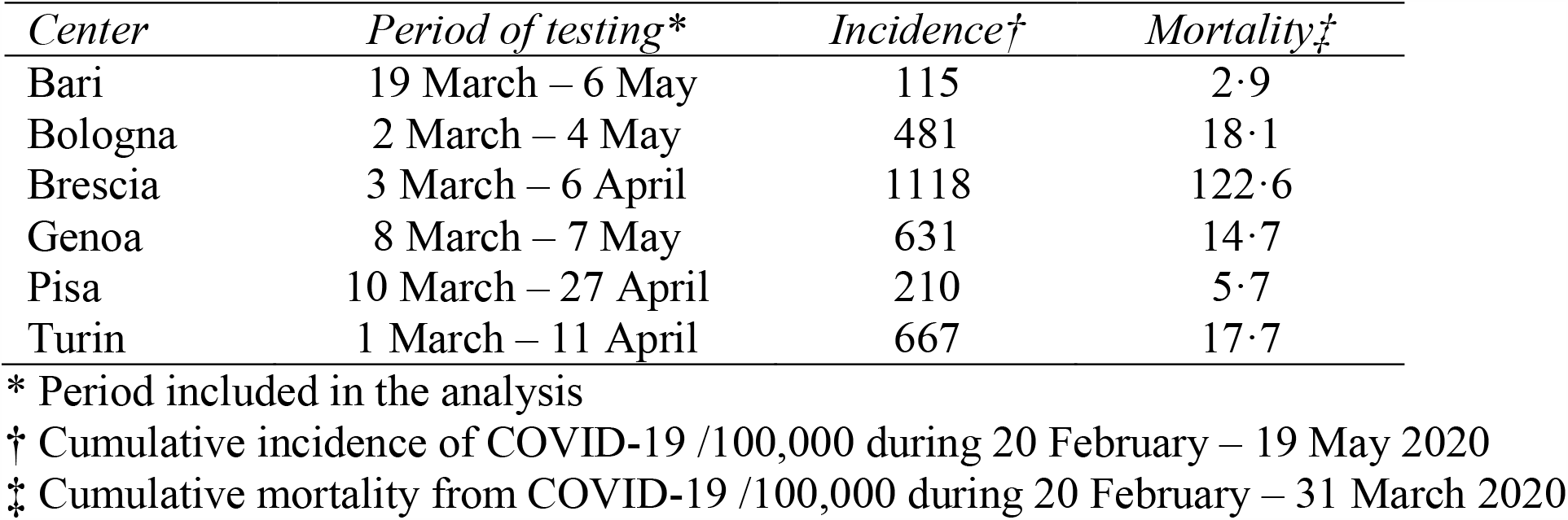
Selected characteristics of surveillance programs and occurrence of COVID-19, by center·

**Table 2.**
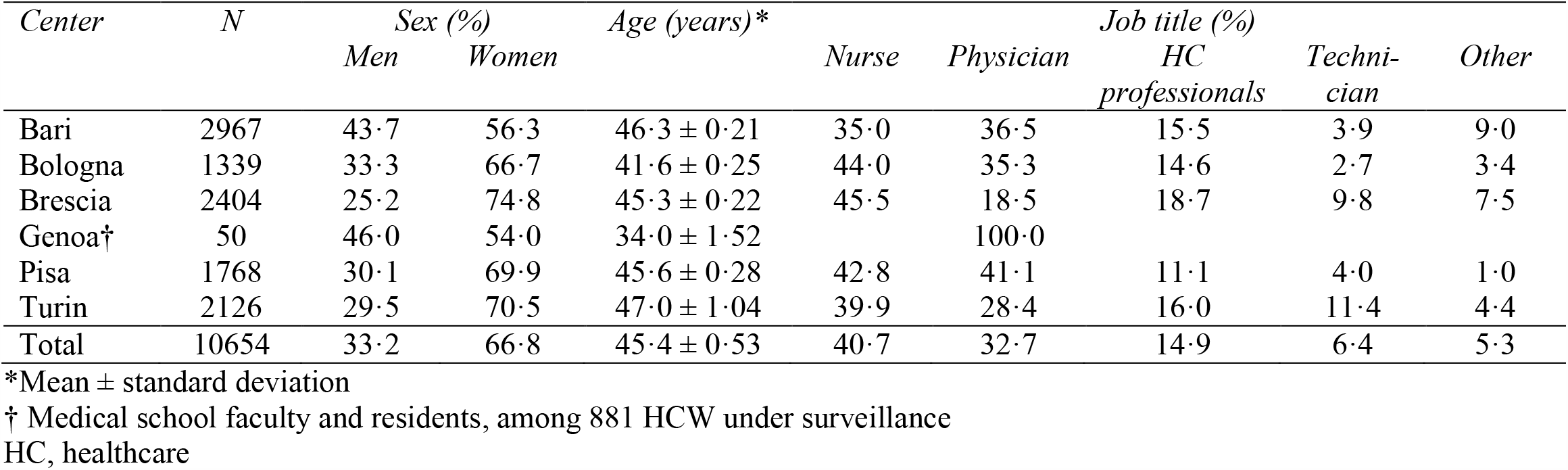
Selected characteristics of study population

Overall, 843 (10.3%) HCW tested positive among 8,203 subjects included in surveillance systems. With the exclusion of the center in Genoa, in which only faculty members and residents who presented a clinical picture suggestive for COVID-19 (suspected cases) were tested (34% positive), the prevalence of HCW positive for SARS-CoV-2 ranged from 3% in Bari and Pisa to 22% in Brescia, while this prevalence in the other centers was between 5% and 7% (Table 3): There was a strong correlation between prevalence of infection in HCW and COVID-19 incidence (r = 0.93, p = 0.002) and mortality (r = 0.99, p = 0.0001) across the study centers.

**Table 3.** Prevalence of SARS-CoV-2 infection among HCW, by center

| *Center* | *N total* | *N positive* | *% positive* |
| --- | --- | --- | --- |
| Bari* | 516 | 14 | 2.7 |
| Bologna | 1339 | 78 | 5.8 |
| Brescia | 2404 | 535 | 22.2 |
| Genoa† | 50 | 17 | 34.0 |
| Pisa | 1768 | 56 | 3.2 |
| Turin | 2126 | 143 | 6.7 |
| Total | 8203 | 843 | 10.3 |

\* Excluding 2451 HCW and subcontract workers who were tested outside high-risk protocol (12 positive results; 0.5%)
† Medical school faculty and residents, among 881 HCW under surveillance

The ORs of SARS-CoV-2 infection for sex, age, and self-reported symptoms are reported in Table 4. Female seemed less at risk of infection than male HCWs; no differences were detected according to age. Self-reported (or measured) fever was strongly associated with infection; additional symptoms associated with SARS-CoV-2 infection include cough, dyspnea, malaise, and ageusia or anosmia; information on the latter two symptoms was available only for a subset of HCW, but they showed a very strong association with infection. Conversely, self-reported sore throat and diarrhea were not associated with infection in this population.

**Table 4.** Odds ratio of SARS-CoV-2 infection according to sex, age, and presence of symptoms

| *Variable* | *N pos/neg* | *OR* | *95% CI* |
| --- | --- | --- | --- |
| Sex * |  |  |  |
| Male | 270/3259 | 1.0 | Ref |
| Female | 584/6528 | 0.79 | 0.67-0.94 |
| Age † |  |  |  |
| <35 | 194/2299 | 1.0 | Ref. |
| 35-44 | 154/1828 | 0.96 | 0.77-1.22 |
| ≥45 | 469/5156 | 1.08 | 0.90-1.30 |
| Symptoms ‡ |  |  |  |
| Fever | 165/179 | 7.10 | 5.48-9.21 |
| Cough | 245/875 | 2.41 | 1.97-2.94 |
| Dyspnea | 38/127 | 1.49 | 1.00-2.21 |
| Sore throat | 123/548 | 0.79 | 0.62-1.01 |
| Ageusia/anosmia | 166/63 | 18.4 | 13.3-25.5 |
| Diarrhea | 7/58 | 0.39 | 0.18-0.87 |
| Malaise | 212/505 | 2.60 | 2.05-3.29 |
| Fever, cough or dyspnea | 317/1214 | 2.82 | 2.32-3.43 |

OR, odds ratio; CI, confidence interval; Ref, reference category
\* OR adjusted for age and center
† OR adjusted for sex and center
‡ Self-reported symptom at least one contact; OR adjusted for sex, age and center

Results on the association between SARS-CoV-2 infection and job-related circumstances of exposure are reported in Table 5. No differences in the risk of infection were detected for job titles, although there was some heterogeneity in results among centers (not shown in detail). Working in a COVID-19 designated department was not a risk factor for infection The analysis on the potential source of infection indicated that contact with a patient was associated with a higher risk of SARS-CoV-2 infection compared to contact with a colleague, which represented the majority of contacts at the workplace.

**Table 5.** Odds ratio of SARS-CoV-2 infection in HCW according to source of contact and use of PPE

| *Variable* | *N pos/neg* | *OR* | *95% CI* |
| --- | --- | --- | --- |
| Job * |  |  |  |
| Nurse | 278/3635 | 1.0 | Ref. |
| Physician | 168/2949 | 1.01 | 0.80-1.26 |
| Health care assistant | 112/1359 | 1.10 | 0.86-1.41 |
| Technician | 55/567 | 1.05 | 0.76-1.45 |
| Other | 36/498 | 0.92 | 0.62-1.36 |
| Department of employment ** |  |  |  |
| Non-COVID-19 designated | 419/7905 | 1.0 | Ref. |
| COVID-19 designated | 144/1185 | 0.96 | 0.76-1.23 |
| Exposure † |  |  |  |
| Out of workplace | 30/138 | 0.98 | 0.47-2.05 |
| At workplace | 350/2706 | 0.52 | 0.21-1.27 |
| Colleague | 223/2715 | 0.60 | 0.43-0.84 |
| Patient | 229/2434 | 1.18 | 0.84-1.65 |
| Use of PPE ‡ |  |  |  |
| Any mask | 267/4794 | 0.69 | 0.47-1.00 |
| Face shield | 121/3164 | 1.22 | 0.83-1.79 |
| Gloves | 215/4053 | 0.72 | 0.50-1.02 |
| Gown | 127/3220 | 1.39 | 0.94-2.09 |
| Any PPE | 317/6427 | 0.94 | 0.76-1.16 |
| Use of mask by contact |  |  |  |
| Any mask (contact) § | 32/742 | 0.52 | 0.32-0.85 |
| Any mask (HCW and contact) | 22/659 | 0.30 | 0.16-0.55 |

OR, odds ratio; CI, confidence interval; Ref, reference category
\* OR adjusted for sex, age, and center
\*\* OR adjusted for sex, age, center, and job title
† Self-reported source of contact at least one contact; OR adjusted for sex, age, center and other sources of contacts
‡ Self-reported PPE at least one contact; OR adjusted for sex, age, center, job title and other PPE
§ Self-reported at least one contact; OR adjusted for sex, age, center and use of mask by HCW

The use of surgical mask was associated with a reduced risk of infection, while the use of a filtering facepiece 2 or 3 (FFP2/FFP3) mask did not appear to confer additional protection. Use of gloves was also associated with a reduced risk of infection, while no difference was detected for use of face shield or gown. After adjusting for personal use of mask, the fact that the contact (patient or colleague) wore a mask was associated with a strong reduction in risk of infection (OR 0.52; 95% CI 0.32-0.85). If both the HCW and the contact wore a mask, the risk of infection was strongly reduced (OR 0.31; 95% CI 0.17-0.57) compared to the situation in which neither did (reference case).

The results of the two-stage meta-analyses replicated those of the pooled analyses; for example, the meta-OR for employment as health care attendant was 1.08 (95% CI 0.80-1.36; p-value of test for heterogeneity = 0.73), that for using a medical mask was 0.61 (95% CI 0.41-0.81, p heterogeneity = 0.92).

## Discussion

This analysis revealed that the prevalence of infection in HCW varied across centers, with results collected in centers with comparable protocols ranging from 3.0% to 22.0%, and was strongly correlated with that of the respective geographic areas. These figures fitted the prevalence of infection and mortality from COVID-19 in the general population of the study areas. Despite the limitations in the available data, in particular those from the general population,^9^ these results confirm and quantify the variability in the risk of infection experienced by HCW according to the distribution of SARS-CoV-2 infection in the patient population of the hospitals where they worked. A survey of 1,097 HCW from six hospitals in the Netherlands also reported ample variability between regions.^10^

Middle-aged women in Italy experienced a slightly higher incidence of SARS-CoV-2 infection compared to men, although incidence at older age and the proportion of cases of severe COVID-19 and death across in the whole age range were lower. For example, as of June 3rd, 2020, the female/male ratio in the number of cases in the age range 30-69 was 1.07.^11^ compared to a ratio of 1.02 in the Italian population. In this population of HCW, on the other hand, the risk of infection was lower in women compared to men; however, in the subset of HCW with information on source of contact, no difference in risk was shown according to gender (OR 0.97; 95% CI 0.74-1.27 after adjustment for contact with colleague and patient). This suggests that any difference may be due to circumstance of exposure rather than gender differences. In a study of 72 HCW from Wuhan, China, men were at higher risk of infection compared to women.^6^ Age was not associated with risk of exposure in this population, similar to a small study from Wuhan, China.^6^ These results suggest that differences by age observed in the general population^11^ might be due either to higher opportunity of exposure, or to higher number of tests performed.

Information on symptoms was available for most HCW undergoing testing for SARS-CoV-2 infection. Fever and, to a lesser extent, cough, dyspnea and malaise were the symptoms most strongly associated with infection, while sore throat and diarrhea did not predict a positive results. Information on ageusia and anosmia was collected in a few centers, and only after several weeks of testing: even if they were available on a relatively small number of HCW, these symptoms were strongly predictive of infection.^12^

The analysis by job title and department of employment represents one of the most important contributions of this study. The lack of a clear pattern in risk according to job categories indicates that all HCW, at least those in the centers included in the analysis, were at comparable risk of becoming infected. The lack of information on the denominators, that include also HCW who were not tested because they did not fulfill the criteria initially established by the Italian Ministry of Health^13^ limits our ability to draw conclusions on the absolute risk in the different occupational groups. To address this question, we analyzed a group of HCW from one center (Bari) who were tested outside the protocol based on contact and symptoms. Among 2373 such HCW, 11 cases of SARS-CoV-2 infection were detected, of whom three among 831 nurses, seven among 798 physicians (OR 4.1; 95% CI 1.0-16.4) and one among 381nurses or other health care professionals (OR 0.7; 95% CI 0.1-6.5). The lack of difference in infection prevalence by job is consistent with the results of a study of 1654 HCW from England ^5^. The lack of difference in risk between HCW who worked in designated COVID-19 departments and those who worked in other departments is reassuring as it indicates that working in high-risk environment did not entail a higher risk of infection, probably because of increased awareness and proper use of PPE by the employees. For example, the proportion of HCW employed in COVID-19 departments who reported using surgical mask or FFP2/3 was 87%, compared to 78% of other HCW. Our results are also comparable to those reported in a study of 183 SARS-CoV-2 positive US HCW, among whom the prevalence of infection was comparable between frontline and non-frontline HCW.^14^

The analysis of circumstances of exposure with a COVID-19 case suggested a role for contact with a patient compared to contact with a colleague or a contact outside the workplace.

The effect of PPE use on the risk of becoming infected with SARS-CoV-2 varied according to the device. The use of a surgical mask or a FFP2/3 mask appeared to be the single most effective approach to reduce risk. We found no difference in the effect of using a FFP2/3 instead of using a surgical mask (results not shown in detail). Reported use of gloves was also associated with a reduced risk of infection, although the result did not reach the formal level of statistical significance, while use of face shield or disposable gown was not associated with reduced risk of infection. It is important to note that this information was self-reported by the HCW, and might be subject to some degree of misclassification, which is likely to be non-differential with respect to infection, since it was collected before the HCW knew about their status. Such misclassification would likely reduced the measured protective effect of PPE. An original and important finding of our analysis is the strong protection offered by the use of a surgical or FFP2/3 mask by both the HCW and the patient: the effects of the two devices appear to be additive, with a measured risk of SARS-CoV-2 infection that was less than one third when both were used compared to when neither was. The World Health Organization recommends that HCW caring for inpatient COVID-19 patients wear a medical mask, gown, gloves and eye protection.^15^ Wearing a medical-grade mask is consistent with our findings, there is strong evidence in the literature that use of medical-grade masks protects against viral infection in both outpatient and inpatient settings,^16,17^ including infection with Coronaviruses.^18^ When we searched for randomized trials comparing the protective effects of surgical vs. FFP2/3 masks against Coronaviruses, we did not identified any for novel SARS-CoV-2 causing COVID-19. The debate concerning the comparison of the protection exerted by FFP2/3 masks compared to surgical masks is still ongoing and needs further research.^19,20^ No data were available in the literature on the effect of mask wearing by the source of exposure (patient or colleague). The evidence concerning which characteristics of other PPE affect their effectiveness in preventing SARS-CoV-2 infection is still limited.^17,21^

Our study suffers from some limitations. Data were collected independently in the different centers; the harmonization process might have generated some misclassification, which however was likely non-differential with respect to infection status since information on risk factors was collected before tests were conducted. Although standard protocols were established at the national level to test HCW, it is possible that some individuals were tested outside such protocols. In addition, rhinopharyngeal swabs detect the presence of SARS-CoV-2 at the time of the test, and it is possible that some HCW who tested negative had been previously infected and developed an asymptomatic disease. In addition, the test itself has estimated sensitivity of 80% or less, while the specificity is as high as 99%.^22^ Finally, no information was available on the viral load of positive tests.

Advantages of our study include, in addition to the large number of HCW and the heterogeneity of their jobs and exposure circumstances, the availability of data on multiple potential determinants of SARS-CoV-2 infection, and the prospective collection of information, that reduces the likelihood of reporting bias.

In conclusion, our study showed the importance of studying SARS-CoV-2 infection in HCW with potential high exposure to the virus, and identified several determinants of infection that are also relevant to other occupational groups and the population at large. More large-scale studies are warranted on HCW from other countries which experienced a range of severity of the COVID-19 epidemic and implemented different approaches for the prevention of infection in HCW.

## Methods

We pooled the data on SARS-CoV-2 infection collected by the occupational medicine centers involved in the surveillance of HCW in large urban Italian hospitals in Bari, Bologna, Brescia, Genoa, Pisa and Turin. Starting in March 2020, surveillance systems were established in each center to monitor HCW for possible infection with SARS-CoV-2, including testing them with swabs to detect SARS-CoV-2 RNA by RT-PCR, in Regional Reference Laboratories, and databases were established to monitor and follow them up. No information on viral load was available.

In all centers, HCW were tested for SARS-COV-2 infection using either a rhinopharyngeal or an-oro-pharyngeal swab.^23^ Samples were analyzed according to the guidelines proposed by the World Health Organization.^13^ In general, HCW were tested if matched one of the following case definitions, although some details differed across different centers, and definitions changed slightly over time:

i. patients with severe acute respiratory illness (fever and at least one sign or symptom of respiratory disease, i.e., cough, shortness of breath, and requiring hospitalization) in the absence of an alternative diagnosis that fully explains the clinical presentation;
ii. patients with acute respiratory illness (fever and at least one sign/symptom of respiratory disease, e.g., cough, shortness of breath), and a history of travel to or residence in a location reporting community transmission of COVID-19 disease prior to symptom onset;
iii. subjects with or without acute respiratory illness or symptoms, who have been in contact with a confirmed or probable COVID-19 case in the absence of adequate protection.

Tests were repeated for most subjects with a positive result to monitor the infection; a proportion of subjects with a negative result were re-tested because of repeated contact with a COVID-19 case. In one center (Bari) additional HCW, including subcontract workers, who did not fulfill the inclusion criteria described above, were also tested. In another center (Genoa) only physicians (faculty members and residents) who presented a clinical picture suggestive for COVID-19 (suspected cases) were tested. These subjects were excluded from the analysis of prevalence of SARS-CoV-2 infection, but were retained in the analysis of determinants of infection. The period during which tests included in this analysis were performed, as well as key characteristics of the COVID-19 epidemic in the provinces (intermediate administrative units) where the centers are located.

HCW in centers located in heavily affected areas (Bologna, Brescia, Genoa, Turin) started to be tested at the beginning of March 2020, while testing started in subsequent weeks in centers located in less affected areas (Bari, Pisa). The present analysis included results of tests performed up to April or early May 2020, depending on center.

Different formats were used to collect data in each center, we established a minimum data record whit this set of variables: basic demographic data, job title, hospital department or department of employment, including working in a designated COVID-19 department, self-reported circumstances of contact with a case, self-reported use of personal protection equipment (PPE), including, in one center, use of PPE by the contact person, and selected self-reported (or measured) symptoms. The list of variables is specified in Supplementary Table 1.

We retained in the analysis HCW with at least one valid test result; the outcome of the analysis was the presence of at least one positive result. Multivariable logistic regression models were fitted to the data to estimate odds ratios (OR) of positive result in at least one test, together with 95% confidence intervals (CI). All models included sex, age group and center as potential confounders. Models including additional potential confounders were also run, but in general there was little evidence of reciprocal confounding between exposure variables. In a secondary analysis, center-specific OR for selected risk factors were combined using a random-effects meta-analysis ^24^ to assess the validity of the data pooling approach.

The study was reviewed and considered exempt by the Ethics Committee of the University of Bologna.

## Data Availability

Primary data have been uploaded on Dryad

https://datadryad.org/stash/dashboard

## Working Group members

### Bari

Luigi De Maria, MD^7^, Antonio Caputi, MD^7^, Stefania Sponselli, MD^7^

### Bologna

Carmine Mastrippolito, MD^2^, Carlotta Zunarelli, MD^2^, Giulia Di Felice, MD^2^, Giovanni Visci, MD^2^

### Brescia

Elisa Albini, MD^10^, Emanuele Sansone, MD^5^, Cesare Tomasi, MS^5^, Andrea Bisioli, MD^5^, Lorenzo Cipriani, MD^5^, Alessandro De Bellis, MD^5^, Mara Maria Tiraboschi, MD^5^, Emilio Paraggio, MD^5^, Sofia Rubino, MD^5^, Michele Capuzzi, MD^5^

### Genoa

Guglielmo Dini, MD^3,4^, Bianca Bruzzone, MD^4^, Nicoletta Debarbieri, MD^4^, Alfredo Montecucco, MD^3,4^, Andrea Orsi, MD^3,4^, Alborz Rahmani, MD^3,4^, Valentina Ricucci, MD^4^

### Pisa

Giovanni Guglielmi, MD^8^, Leonardo Fiorentino, MD^9^, Cinzia Brilli, MD^8^

### Turin

Alessandro Godono, MD^6^, Michael Declementi, MD^6^, Ihab Mansour, MD^6^, Nicolò Milanesio, MD^6^, Giacomo Garzaro, MD^6^, Antonio Scarmozzino, MD^11^, Attilia Gullino, MD^11^

## Contribution of the authors

PB, FV, PA, PD, GS: design of the study

FV, PD, LV, GDP, ACr, EPi: supervision of data collection

GD, ESal, MCo, ST, LDM, ACa, SS, CM, CZ, GDF, GV, EA, ESan, CT, AB, LC, ADB, MMT, EPa, SR, MCa, GI, BB, ND, AM, AR, AO, VR, AG, MD, IM, NM, GGa, AS, GGu, VG, LF, CB: data collection

GI: coordination of laboratory testing of SARS-CoV-2

PB, FV, LV, GDP, ACr, EPi, ES, CT, IM: data harmonization, statistical analysis

PB: drafting of the manuscript

PB, FV, PA, PD, GS, LV, GDP, ACr, EPi, GD, GI: review of each phase of the study

All authors: review and approval of the manuscript

## Declarations

a. All methods were carried out in accordance with relevant guidelines and regulations.
b. The study was reviewed and considered exempt by the Ethics Committee of the University of Bologna.
c. Informed consent was not deemed necessary by the Ethics Committee of the University of Bologna.

---

**Supplementary Table 1.**
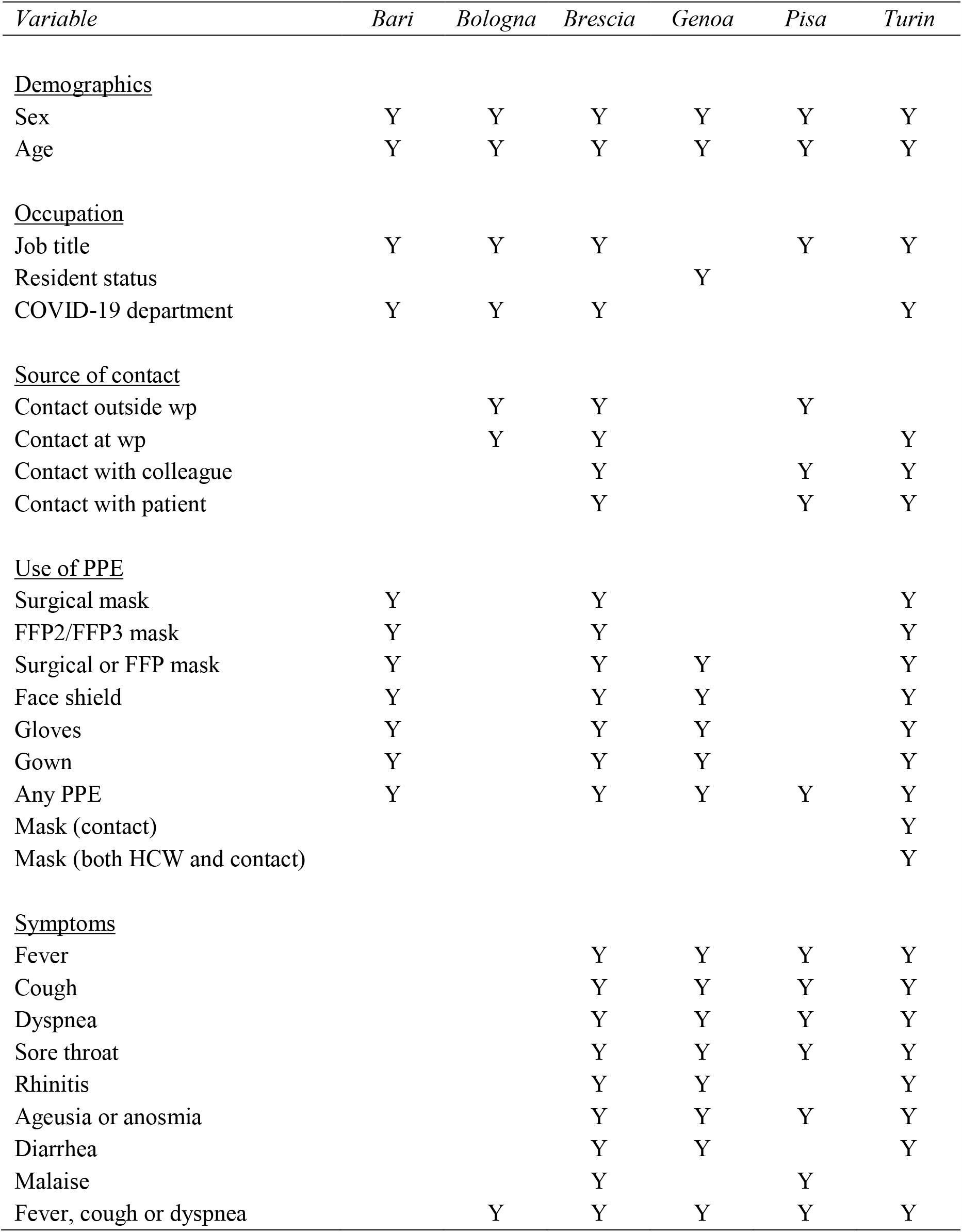
Availability of information by center

| *Variable* | *Bari* | *Bologna* | *Brescia* | *Genoa* | *Pisa* | *Turin* |
| --- | --- | --- | --- | --- | --- | --- |
| <u>Demographics</u> |  |  |  |  |  |  |
| Sex | Y | Y | Y | Y | Y | Y |
| Age | Y | Y | Y | Y | Y | Y |
| <u>Occupation</u> |  |  |  |  |  |  |
| Job title | Y | Y | Y |  | Y | Y |
| Resident status |  |  |  | Y |  |  |
| COVID-19 department | Y | Y | Y |  |  | Y |
| <u>Source of contact</u> |  |  |  |  |  |  |
| Contact outside wp |  | Y | Y |  | Y |  |
| Contact at wp |  | Y | Y |  |  | Y |
| Contact with colleague |  |  | Y |  | Y | Y |
| Contact with patient |  |  | Y |  | Y | Y |
| <u>Use of PPE</u> |  |  |  |  |  |  |
| Surgical mask | Y |  | Y |  |  | Y |
| FFP2/FFP3 mask | Y |  | Y |  |  | Y |
| Surgical or FFP mask | Y |  | Y | Y |  | Y |
| Face shield | Y |  | Y | Y |  | Y |
| Gloves | Y |  | Y | Y |  | Y |
| Gown | Y |  | Y | Y |  | Y |
| Any PPE | Y |  | Y | Y | Y | Y |
| Mask (contact) |  |  |  |  |  | Y |
| Mask (both HCW and contact) |  |  |  |  |  | Y |
| <u>Symptoms</u> |  |  |  |  |  |  |
| Fever |  |  | Y | Y | Y | Y |
| Cough |  |  | Y | Y | Y | Y |
| Dyspnea |  |  | Y | Y | Y | Y |
| Sore throat |  |  | Y | Y | Y | Y |
| Rhinitis |  |  | Y | Y |  | Y |
| Ageusia or anosmia |  |  | Y | Y | Y | Y |
| Diarrhea |  |  | Y | Y |  | Y |
| Malaise |  |  | Y |  | Y |  |
| Fever, cough or dyspnea |  | Y | Y | Y | Y | Y |

